# Metabolomic Signatures of Brain Atrophy and Ibudilast Response in Progressive Multiple Sclerosis

**DOI:** 10.64898/2026.05.21.26353780

**Authors:** Mingjing Chen, Rose Noroozi, Matthew D. Smith, Muraleetharan Sanjayan, Cesar Higgins Tejera, Pavan Bhargava, Blake E. Dewey, Ellen M. Mowry, Kathryn C. Fitzgerald

**Author notes:** **Correspondence to**: Kathryn C. Fitzgerald, ScD. Department of Neurology, Johns Hopkins University School of Medicine Baltimore, MD, USA, 33612.

## Abstract

**Background:** Progressive multiple sclerosis (MS) is characterized by ongoing neurodegeneration and limited therapeutic options. Circulating metabolites provide insight into disease biology, yet biomarkers that predict disability progression and reflect treatment response are lacking. We aimed to identify metabolomic signatures associated with longitudinal MRI measures of brain atrophy and to evaluate whether ibudilast treatment was associated with metabolite trajectories over time.

**Methods:** We repeatedly profiled 1,726 plasma metabolites using untargeted UPLC-MS/MS in 244 participants (mean age 55.6 years; 53.3% female; 3.3% non-White) from the 96-week SPRINT-MS randomized trial of oral ibudilast (≤100 mg daily; n=123) versus placebo (n=121). Weighted gene co-expression network analysis was used to derive groups of related metabolites. Associations between baseline metabolites groups and longitudinal MRI outcomes were evaluated using linear mixed-effects models adjusted for demographic, clinical, and treatment covariates. The primary outcome was the rate of whole-brain atrophy measured by brain parenchymal fraction (BPF), defined as the proportion of intracranial volume occupied by brain tissue. Secondary outcomes included white matter fraction (WMF), gray matter fraction (GMF), and cortical thickness (CTH). Metabolite groups nominally associated with MRI outcomes (p<0.05) were followed by individual metabolite analyses to identify potential drivers. Significant metabolites were tested for replication in a comparable real-world observational HEAL-MS cohort with longitudinal MRI data (n=249; mean age 56.3 years; 71.1% female; 19.4% non-White). Lastly, we tested whether ibudilast treatment was associated with metabolite trajectories and performed metabolite set enrichment analysis.

**Findings:** Higher baseline levels of glycerophospholipids were associated with slower decline in both BPF and WMF, and sphingomyelins were similarly associated with slower BPF decline. For example, higher 1-palmityl-2-stearoyl-GPC (O-16:0/18:0) levels were associated with slower BPF decline in SPRINT-MS (β=0.016 [0.008, 0.024]; p=4.35×10⁻^5^) and replicated in HEAL-MS (β=0.108 [0.006, 0.211], p=3.90×10⁻^2^). Metabolites associated with GMF preservation were enriched in androgenic steroids and steroid sulfates, with consistent positive associations observed in the replication cohort, whereas metabolites inversely associated with CTH were predominantly xenobiotic-related. Ibudilast treatment was associated with increased sphingomyelin species (e.g., palmitoyl sphingomyelin (d18:1/16:0); β = 0.185 [0.085, 0.286], FDR = 1.79×10^−2^) and decreased levels of amino acid-related metabolites (e.g., anthranilate; β = –0.270 [–0.403, –0.137]; FDR = 3.87×10^−2^). Pathway-based analyses corroborated these findings, highlighting glycerophospholipid and sphingolipid metabolism as key pathways implicated in brain atrophy in MS.

**Interpretation:** Distinct lipid subsets were associated with slower brain atrophy in people with MS, and ibudilast treatment was associated with metabolite alterations in potentially neuroprotective directions. Metabolomics may provide prognostic and pharmacodynamic biomarkers for progressive MS.

**Funding:** The study was supported by the National Institute of Neurological Disorders and Stroke (NINDS) grant R01NS133005 and the National Institute of Nursing Research (NINR) grants R01NR018851.

**Research in context:** *Evidence before this study:* Circulating metabolites are altered in people with multiple sclerosis (MS). Before conducting this work, we systematically searched PubMed from database inception to March 23, 2026, for articles published in English using the search terms (“metabolomics” OR “plasma metabolites”) AND (“multiple sclerosis” OR “MS”) AND (“MRI” OR “brain atrophy” OR “brain parenchymal fraction” OR “gray matter” OR “white matter”) AND (“longitudinal” OR “progression” OR “brain atrophy”). We also reviewed reference lists of relevant publications. Prior studies have linked selected metabolites to clinical disability and brain atrophy in MS. However, most studies have been cross-sectional, limited by small sample sizes, or focused on case-control comparisons. Importantly, few studies have evaluated longitudinal associations between circulating metabolites and MRI-derived measures of brain atrophy, and studies integrating clinical trial data, external replication, and treatment-related metabolic changes remain scarce.

*Added value of this study:* In a multicenter randomized clinical trial with longitudinal metabolomic profiling and MRI outcomes, we identified lipid-related metabolic signatures associated with brain atrophy, with consistent directionality observed in an independent cohort. We further demonstrated that ibudilast treatment was associated with longitudinal changes in specific metabolites, linking metabolic pathways to both disease progression and therapeutic response.

*Implications of all the available evidence:* These findings support circulating metabolomic signatures as potential markers of brain atrophy in MS. Metabolomics may provide a scalable approach to identify individuals at risk of progressive brain tissue loss and to inform future mechanistic and therapeutic investigations targeting metabolic pathways involved in disability progression.

## Introduction

Multiple sclerosis (MS) is a chronic inflammatory and neurodegenerative disease of the central nervous system (CNS) affecting nearly 2.8 million individuals worldwide.^1^ It is characterized by demyelination in both white and gray matter, leading to axonal loss, brain atrophy, and irreversible disability.^2,3^ Quantitative MRI-derived measures, including brain parenchymal fraction (BPF), gray matter fraction (GMF), white matter fraction (WMF), and cortical thickness (CTH) are well-established markers of brain atrophy and are strongly associated with long-term clinical outcomes in MS.^4–8^ BPF, in particular, is widely used in clinical trials as a measure of global brain atrophy,^5,9–11^ while GMF and CTH capture gray matter-specific neuroaxonal loss, which more closely correlates with disability progression.^11–14^ WMF provides complementary information on white matter structural integrity, enabling assessment of compartment-specific neurodegenerative processes.^12,15^ Despite these strengths, MRI primarily reflects accumulated structural injury and may not directly capture the dynamic biological processes that drive tissue loss. Identifying complementary biofluid biomarkers could improve risk prediction, provide insight into mechanisms of tissue loss, and enhance monitoring of brain atrophy and treatment effects, especially as it relates to progressive MS.

Metabolomics offers a high-throughput approach to quantify small metabolites (<1500 Da) that reflect real-time biochemical activity and integrated effects of genetic, environmental, and therapeutic influences. In MS, metabolic pathways including lipid metabolism, mitochondrial function, and amino acid turnover are closely linked to myelin integrity, neuroaxonal injury, and immune regulation.^16–19^ Previous studies,^16–19^ including work from our group^8,20^ have reported metabolomic alterations in people with MS (pwMS) across lipid, amino acid, nucleotide, and energy metabolism pathways.^16–18,20–28^ Some metabolites have been associated with clinical disability and brain atrophy.^4,5,20^ However, most studies have been cross-sectional, limited by small sample sizes, or focused on case-control comparisons. Critically, the longitudinal relationships between circulating metabolites and brain atrophy, and their potential modulation by therapy, remain poorly understood.

Longitudinal clinical trial cohorts with standardized imaging and treatment exposure provide a rigorous framework to address these gaps. The SPRINT-MS trial, a multicenter phase II randomized study of ibudilast in progressive MS, demonstrated a reduction in the rate of brain atrophy compared with placebo.^9,29^ Leveraging this well-characterized cohort, we performed large-scale untargeted plasma metabolomic profiling in 244 participants with up to 96 weeks of follow-up and evaluated key findings in an independent replication cohort of 249 individuals with MS. We aimed to (1) determine whether baseline circulating metabolites are associated with longitudinal MRI measures of brain atrophy, independent of clinical covariates, and (2) evaluate whether longitudinal metabolite trajectories are associated with ibudilast treatment. In addition, we performed pathway enrichment analyses to identify metabolic pathways underlying metabolites associated with the MRI measures of brain atrophy.

## Methods

### An overview of the methodology employed in this paper can be found in Figure 1. Study population

We conducted a multi-stage analysis using untargeted metabolomics and longitudinal MRI data from the SPRINT-MS randomized clinical trial (discovery cohort) and the independent Home-based Evaluation of Actigraphy to Predict Longitudinal Function in MS (HEAL-MS) observational cohort (replication cohort). In SPRINT-MS, we performed weighted gene co-expression network analysis (WGCNA) of metabolite data, prospective evaluations of baseline metabolites in relation to longitudinal MRI-derived measures of brain atrophy, longitudinal treatment-response analyses of metabolite trajectories, and pathway enrichment analyses.

**Figure 1.**
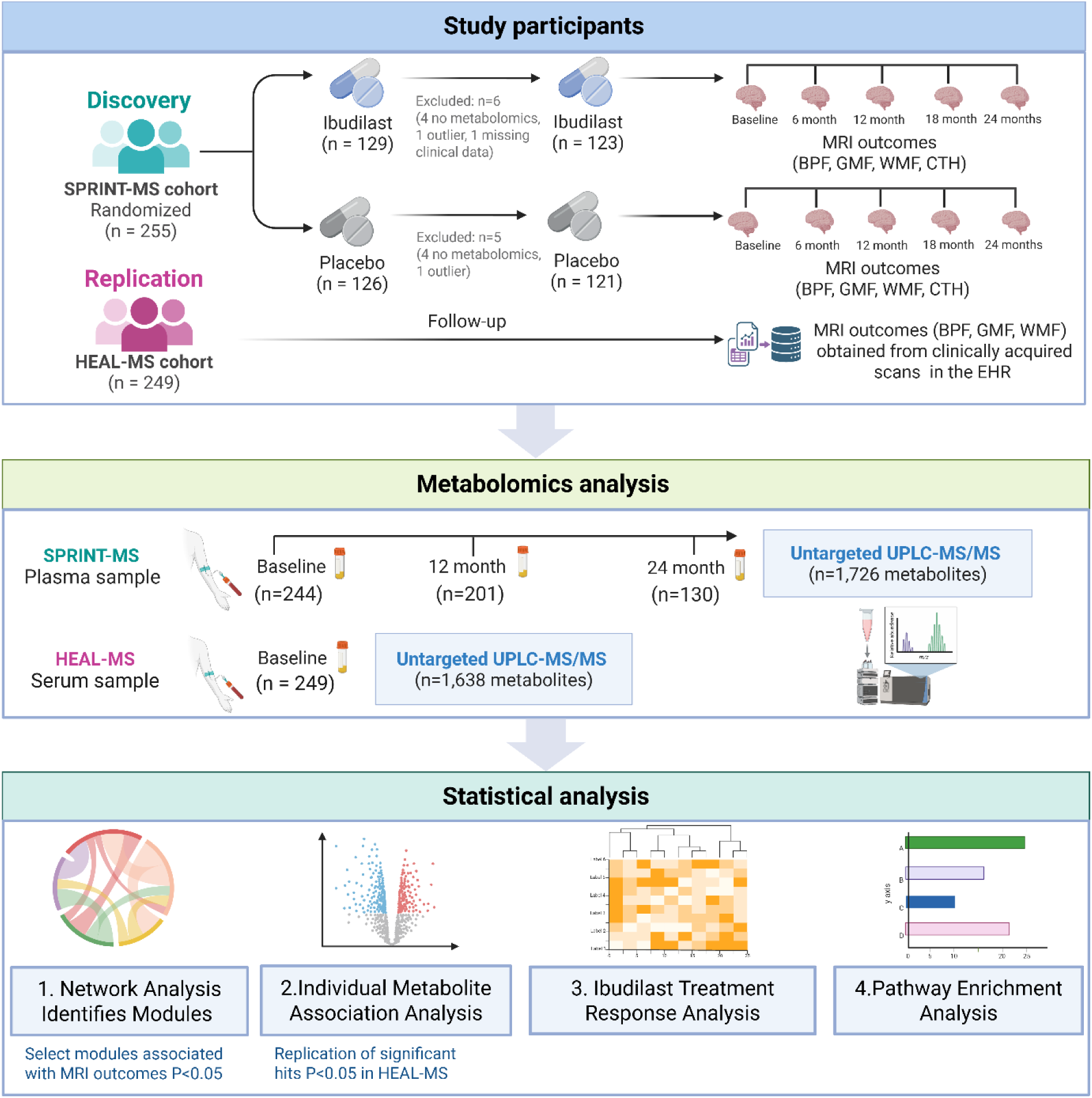
Overview of the study design and workflow. The discovery phase used the SPRINT-MS randomized clinical trial (n=244) to evaluate associations between circulating metabolites and longitudinal MRI-derived measures of brain atrophy and to assess treatment-related changes in metabolite trajectories. The replication phase used a comparable real-world observational HEAL-MS cohort (n=249) to validate prospective associations between baseline metabolites and subsequent MRI outcomes. Untargeted metabolomics quantified 1,726 metabolites in longitudinal plasma samples from SPRINT-MS and 1,638 metabolites in baseline serum samples from HEAL-MS. In SPRINT-MS, weighted gene co-expression network analysis identified metabolite modules associated with MRI trajectories. Individual metabolite-level associations were then evaluated prospectively. Significant baseline metabolite-MRI associations were tested for replication in HEAL-MS. Pathway enrichment analysis was performed in SPRINT-MS to characterize biological pathways represented by MRI-associated metabolites.

Prospective associations between baseline metabolites and subsequent brain atrophy identified in SPRINT-MS were then evaluated for replication in HEAL-MS.

### The SPRINT-MS cohort (Discovery)

SPRINT-MS is a phase II, multicenter, randomized, placebo-controlled clinical trial of oral ibudilast, a phosphodiesterase inhibitor with anti-inflammatory and neuroprotective properties that modulates microglial activation and cytokine signaling, administered at doses up to 100 mg daily over 96 weeks. The study demonstrated the benefit of ibudilast on the primary endpoint, the rate of brain atrophy as measured by BPF. The study design, detailed protocol, primary and secondary results have been described previously.^9,29^ Briefly, participants were randomized 1:1 to ibudilast or placebo across 28 study sites, stratified by disease subtype (primary progressive MS [PPMS] vs secondary progressive MS [SPMS]) and current disease-modifying therapy (DMT) use (glatiramer acetate or interferon-β vs none). Eligible participants were 21-65 years old, had Expanded Disability Status Scale (EDSS) scores between 3.0 and 6.5, and demonstrated disease progression within the previous two years. Key exclusion criteria included recent relapse, systemic glucocorticoid use within three months of screening, use of DMT other than interferon-β or glatiramer acetate, concurrent medications interfering with ibudilast, current depression, or inability to undergo MRI. A total of 255 eligible participants were initially enrolled. We included a subset of 244 participants (96%) with available baseline plasma samples and complete metabolomics data. Participants provided blood samples approximately annually.

### The HEAL-MS cohort (Replication)

HEAL-MS is a longitudinal observational cohort of pwMS conducted at the Johns Hopkins Multiple Sclerosis Precision Medicine Center of Excellence conducted from January 2021 to March 2023 (NIH R01NR018851). Detailed descriptions of the study design and phenotype collection have been published previously.^30^ Briefly, the cohort enrolled 255 participants, including 170 with relapsing-remitting MS and 85 with progressive MS, matched on age, sex, race, and DMT use. Eligible participants were aged ≥ 40 years, had an EDSS score ≤ 6.5, had no relapse within 6 months prior to enrollment, and had no comorbid conditions limiting physical activity (e.g., heart failure, uncontrolled hypertension). To ensure temporal alignment between baseline metabolomics and imaging measures, eligible participants were those for whom baseline MRI was obtained within ±180 days of the blood draw. Of the 255 enrolled participants, 253 completed baseline metabolomic profiling. Four participants were excluded due to the absence of MRI data within the prespecified time window, resulting in a final analytic sample of 249 participants.

### Ethics statement

This study was conducted in accordance with the Declaration of Helsinki. SPRINT-MS was approved by the institutional review boards of all participating institutions and registered at <u>ClinicalTrials.gov</u> (NCT01982942). HEAL-MS was approved by the Johns Hopkins University institutional review board (IRB00243681). All participants in both studies provided written informed consent. All data are de-identified to the greatest extent possible.

### Assessments of MS neurological imaging outcomes

In SPRINT-MS, MRI was performed at baseline and at weeks 24, 48, 72, and 96 using standardized acquisition protocols^31–35^ across all study sites. All scans were obtained on 3T MRI systems (Siemens Trio/Prisma or Skyra; General Electric 12X or higher). Image acquisition protocols included (1) 3D spoiled gradient-recalled echo; (2) proton density weighted and T2 weighted FLAIR; (3) 3D spoiled-gradient recalled echo with selective excitation with and without magnetization transfer pulse; and (4) 64 direction high angular resolution diffusion imaging. Each site used its scanner’s default settings for magnetization transfer pulses, and all MRI data were processed centrally to ensure consistency across scanners and time points.

The primary MRI outcome was the rate of whole-brain atrophy, quantified by the brain parenchymal fraction (BPF), defined as the proportion of intracranial volume occupied by brain tissue and normalized for head size, derived using a standardized MRI processing pipeline.^36,37^ Secondary MRI outcomes included: 1) white matter fraction (WMF): the proportion of intracranial volume occupied by white matter, reflecting white matter loss; 2) gray matter fraction (GMF): the proportion of intracranial volume occupied by cortical and deep gray matter; 3) cortical thickness (CTH): the average thickness of the cortical mantle derived from automated surface-based morphometry. Detailed MRI image acquisition and quality assurance have been described previously.^9,29^ Safety outcomes were assessed by site investigators, who recorded adverse and serious adverse events at each study visit. All serious adverse events were independently reviewed by a medical monitor.

For HEAL-MS, research brain MRI scans were performed on a 3T Siemens system at study entry and again at the Year 2 visit. When available, interim MRIs acquired during routine clinical care using the same protocol were incorporated into the longitudinal dataset.^38,39^ Images were processed using a standardized longitudinal pipeline optimized for multi-site and clinically acquired data.^40,41^ This included image harmonization, lesion segmentation, regional brain segmentation, cortical surface reconstruction, and normalization to intracranial volume. The processing framework has been described in detail previously.^42–44^

### Assessment of clinical covariates

In both cohorts, demographic information (age, sex, and race), lifestyle (smoking status), medical history, and prescription medication use were abstracted from electronic health records as previously described.^9,29^ Body mass index (BMI) was calculated as body weight in kilograms divided by the square of height in meters. Participants self-identified their race as White, Black, or Other race. Smoking status was categorized as never, former, or current. Disease-modifying therapy (DMT) use at baseline was classified as treated or untreated. Among treated participants, DMTs were further categorized as efficacy level into low-efficacy therapies (injectable agents, including interferon-beta and glatiramer acetate), moderate-efficacy therapies (oral agents, including teriflunomide, sphingosine-1-phosphate receptor modulators, and fumaric acid esters), and high-efficacy therapies (infusion or immune reconstitution therapies, including anti-CD20 agents, natalizumab, alemtuzumab, and cladribine). Disease duration was defined as the number of days since MS diagnosis. In SPRINT-MS, participants were limited to progressive MS and classified as primary progressive MS (PPMS) or secondary progressive MS (SPMS) according to trial eligibility criteria. In HEAL-MS, participants were categorized as relapsing-remitting MS or progressive MS based on treating neurologist assessment.

### Metabolomic data acquisition, pre-processing, and quality control

In SPRINT-MS, blood samples were collected and processed according to a standardized protocol and shipped to a centralized repository for storage at -80°C. A total of 1,745 metabolites (including 1,409 named biochemicals) were quantified from 579 plasma samples collected at baseline (n=246), 12 months (n=202), and 24 months (n=130) using ultrahigh-performance liquid chromatography-tandem mass spectrometry (UPLC-MS/MS) at Metabolon, Inc. (Durham, NC). Quality control (QC) samples were analyzed alongside study samples to ensure analytical consistency across batches and runs. Technical reproducibility was confirmed using coefficients of variation (CVs) derived from duplicate samples. Outlier detection was performed using principal component analysis (PCA) and Euclidean distance-based sample connectivity measures.^45,46^ Samples exceeding 3 standard deviations from the mean of PC1 or PC2, or with Z<4 based on connectivity statistics (Zₖ), were excluded. No significant batch effects were observed (**Supplemental Methods**).

After further excluding outlier samples and those with missing covariates, the final analysis included 1,726 metabolites (1,390 identified metabolites) from 575 plasma samples (244 at baseline, 201 at 12 months, 130 at 24 months follow-up) obtained from 244 pwMS (n=123 ibudilast; n=121 placebo).

In HEAL-MS, metabolomic profiling was performed using 1,638 metabolites (including 1,325 named biochemicals) from 249 serum samples collected at baseline by untargeted UPLC-MS/MS at Metabolon, Inc. (Durham, NC). Metabolomic profiling in HEAL-MS was performed using the same untargeted UPLC-MS/MS platform and standardized Metabolon preprocessing procedures as described for SPRINT-MS. Metabolite data in both cohorts were obtained as batch-normalized and imputed values from Metabolon using median scaling and minimum value imputation. Metabolites identified in SPRINT-MS were mapped to those available in HEAL-MS cohort based on shared biochemical annotations. These overlapping metabolites were used to replicate the findings from SPRINT-MS.

### Statistical analysis

Statistical analyses were performed using R version 4.1.1 (R Foundation for Statistical Computing, Vienna, Austria). All continuous variables, including metabolite levels, were standardized to zero mean and unit variance prior to analysis unless otherwise stated. To account for multiple testing, we controlled the false discovery rate (FDR) using the Benjamini–Hochberg procedure,^47^ with FDR-adjusted p values<0.05 considered statistically significant.

### Network analysis

Given the high correlations among plasma metabolites, we performed the Weighted Gene Co-expression Network Analysis (WGCNA)^48^ to 1,726 baseline plasma metabolites in SPRINT-MS. Additional details are described in the **Supplemental methods**. Briefly, this agnostic approach clustered metabolites into modules of highly correlated metabolites (metabolite sets) without requiring prior assumptions. WGCNA facilitates the unbiased construction of correlated metabolite networks and offers a robust framework to identify highly correlated hub metabolites, centrally located within each module, to address the multiple testing challenges associated with analyzing hundreds of metabolites and to investigate metabolic set-wide relationships to derive meaningful biological insights from functionally correlated metabolites. For each significant module (p<0.05), we calculated the module eigenmetabolite (first principal component), which represents the dominant variation of all metabolites in that module. These eigenmetabolites were used as predictors in downstream network analyses.

### Prospective Association Analyses

To evaluate whether baseline metabolites are associated with longitudinal MRI outcomes, we used linear mixed-effects (LME) models with random intercepts to account for repeated measures within participants. We first tested module eigenmetabolites derived from WGCNA as predictors of each repeated MRI outcome (BPF, WMF, GMF, and CTH). In these models, the independent variable included the baseline module eigenmetabolites, time (defined as days since baseline MRI), and their interaction term (baseline module × time), with the interaction term representing the association between baseline metabolite levels and longitudinal change in MRI outcomes, and the dependent variables were repeated measures of each MRI outcome. All models were adjusted for age, sex, race, BMI, smoking status, treatment group, MS subtype, disease duration, and baseline DMT use, and were fitted using restricted maximum likelihood (REML).

For modules significantly associated with MRI outcomes (p<0.05), we then performed follow-up analyses at the individual metabolite level to identify specific metabolites driving the association. These complementary analyses used the same LME modeling framework and covariate adjustment. Metabolites with nominal significance (p<0.05) in SPRINT-MS were subsequently evaluated in the independent HEAL-MS cohort using the same modeling frameworks and covariate adjustment, excluding treatment group. Replication was defined as metabolites with p<0.05 and consistent directions of association across both cohorts. Finally, results from SPRINT-MS and HEAL-MS were combined using inverse-variance weighted fixed-effects meta-analysis.

### Ibudilast-associated metabolite trajectory analyses in the discovery cohort

To evaluate whether ibudilast affected longitudinal metabolite trajectories, we fitted LME models for each WGCNA module eigenmetabolite as the dependent variable. The independent variables included treatment group (ibudilast vs. placebo), time (days since baseline MRI), and their interaction term (treatment × time), where the interaction term estimates the difference in longitudinal change between ibudilast and placebo (i.e., a drug-associated slope difference). All models included random intercepts to account for within-participant correlation of repeated measures and were adjusted for age, sex, race, BMI, smoking status, disease duration, MS subtype, and DMT use at baseline. We further examined individual metabolites belonging to modules that showed a significant treatment × time effect in the primary analysis (p<0.05) using the same analysis as described above, with FDR correction applied across tested metabolites.

### Pathway enrichment analysis in SPRINT-MS

We performed metabolite set enrichment analysis (MSEA)^49^ to identify metabolic pathways associated with each MRI outcome. More details are described in the **Supplemental methods**. Briefly, for the a *priori*-defined metabolite pathway analysis, enrichment was evaluated using known metabolic pathways . MSEA was conducted on pre-ranked individual metabolite statistics (i.e., the statistic obtained from individual metabolite LME models with outcomes), using curated biological pathways annotations (sub-pathway) obtained from Metabolon, Inc., as previously described.^50^ The enrichment score (ES) indicates the degree of overrepresentation of a metabolite set at the extremes of the entire ranked list of metabolites. To account for variability in metabolite set sizes, the normalized enrichment score (NES) was calculated, enabling meaningful comparisons across pathways. The FDR-adjusted p-value was used to evaluate the statistical significance of NES. Importantly, this pathway-based approach is complementary to our network-based analyses (e.g., WGCNA), which are agnostic to predefined pathway annotations. In contrast to module-restricted analyses, MSEA was performed using all measured known metabolites, without restricting to metabolites within significant modules.

### Sensitivity Analysis

To assess the robustness of our findings, we conducted several sensitivity analyses. First, sex-specific effects were evaluated by including an interaction term (metabolite × sex) in the models. Second, because metabolites associated with GMF were predominantly steroidogenic lipids, we conducted sex-stratified analyses to further examine potential sex-related differences in these associations.

## Results

Baseline characteristics for the discovery (SPRINT-MS, n=244) and replication (HEAL-MS, n=249) cohorts are summarized in **Table 1**. The mean age was comparable between cohorts (55.6 ± 7.3 years in SPRINT-MS; 56.2 ± 9.9 years in HEAL-MS), though the replication cohort was predominantly female (71.1% vs. 53.3% in discovery) and demonstrated greater racial diversity (17.3% Black vs. 2.5% in discovery). Notably, use of DMT was substantially higher in the replication cohort (79.1%) than in the discovery cohort (24.2%). Mean disease duration was approximately 12 years for both groups. Baseline MRI measures were generally consistent across cohorts, with mean BPF values of 0.80 ± 0.03 and 0.78 ± 0.04 in the discovery and replication cohorts, respectively.

**Table 1.**
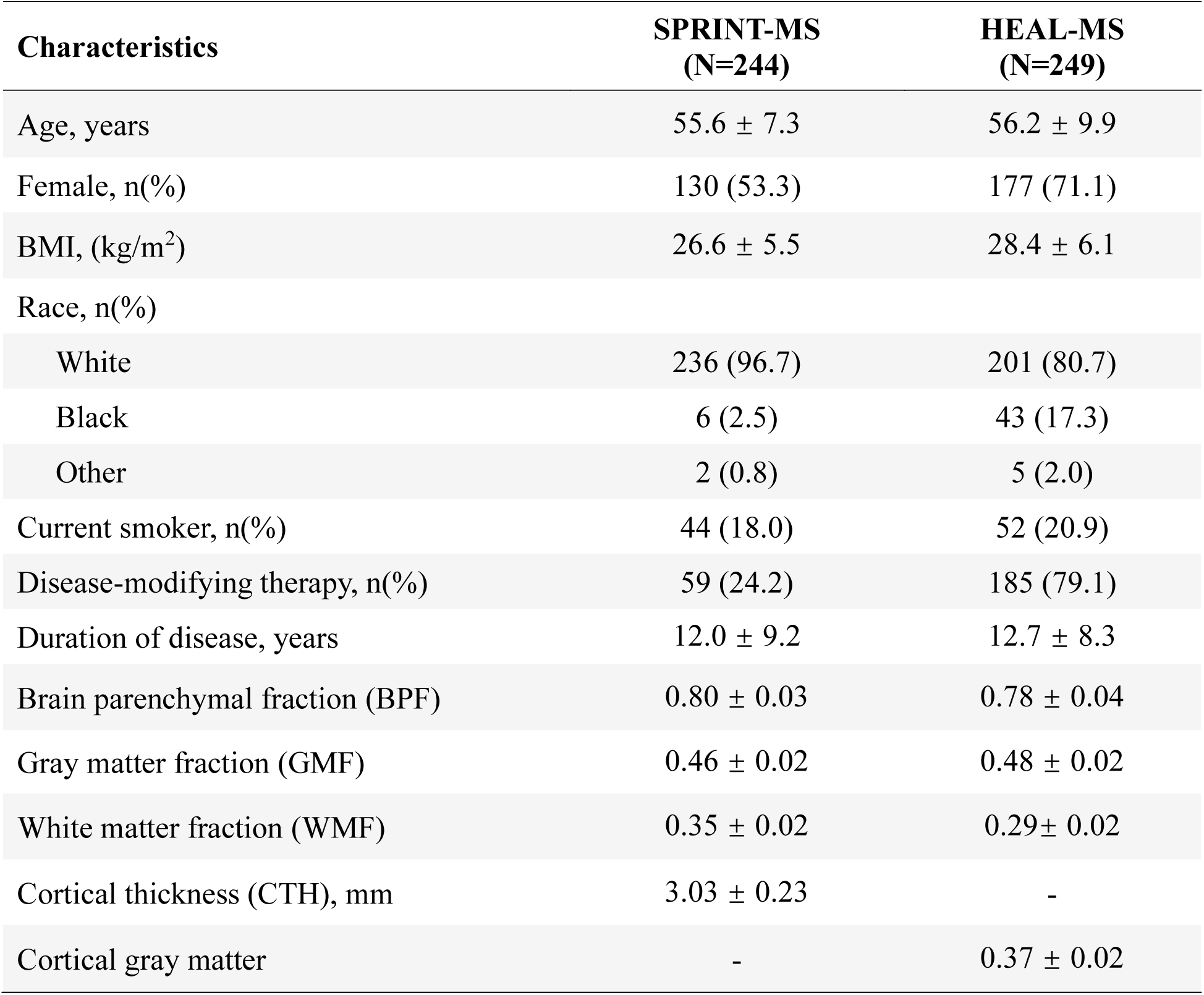
Baseline characteristics of participants in the SPRINT-MS (discovery) and HEAL-MS (replication).

### Prospective associations between baseline metabolites and longitudinal MRI outcomes

In SPRINT-MS, WGCNA classified 1,726 baseline plasma metabolites into 18 modules of correlated metabolites (**Supplemental Table S1**). After adjustment for clinical covariates, 10 modules were associated with at least one longitudinal MRI outcome at p<0.05 and five remained significant following FDR correction (**Table 2**, **Figure 2A**). BPF showed the broadest module-level signal, with seven nominally associated modules. Modules enriched for glycerophospholipids (module 10), fatty acids and acyl lipids (module 9), sphingolipids (module 8), and cofactors/vitamins (module 5) were associated with slower BPF decline (β range: 0.008 [95% CI, 0.001, 0.016] to 0.014 [0.007, 0.022]). In contrast, modules enriched for branched-chain amino acid catabolism (module 6) and xenobiotic/caffeine metabolism (module 2) were associated with faster BPF decline (β range: −0.009 [−0.016 to −0.001] to −0.010 [−0.018 to −0.003]). For WMF, the fatty acid/acyl lipid-enriched module (module 9) was associated with slower WMF decline (β: 0.007 [0.0002, 0.013]). For GMF, modules enriched for branched-chain amino acid metabolism (module 6), androgenic steroids and steroid sulfates (module 4), and xenobiotic/caffeine metabolism (module 2) were associated with slower GMF decline (β range: 0.005 [0.001, 0.009] to 0.006 [0.002, 0.010]). For CTH, modules enriched for gut microbial/aromatic xenobiotic metabolites (module 3) and bile acid metabolism (module 7) were associated with faster CTH decline over time (β range: −0.009 [−0.018 to −0.0001] to −0.013 [−0.022 to −0.005]).

**Figure 2.**
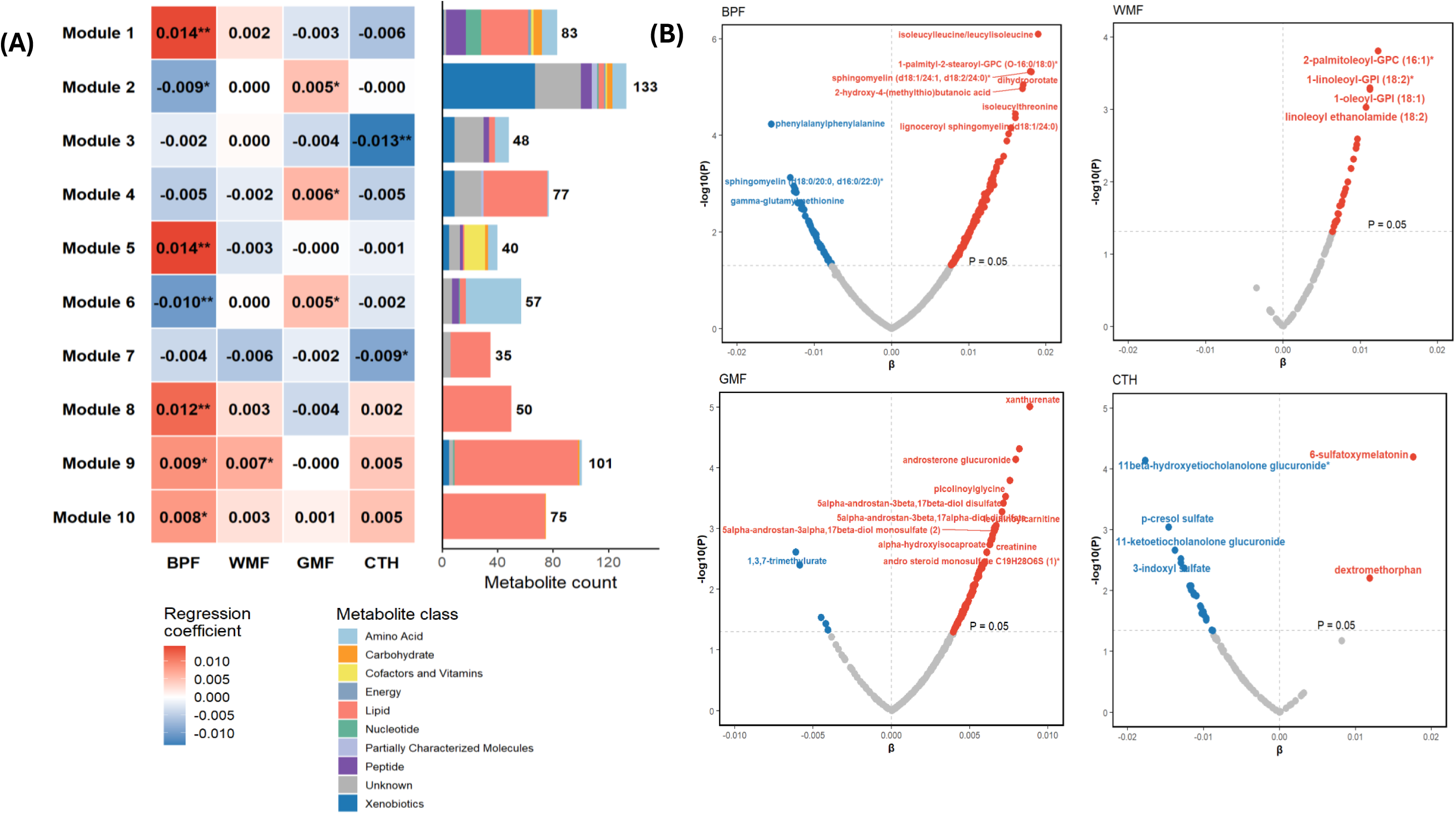
Baseline metabolite modules associated with longitudinal MRI outcomes in SPRINT-MS. (A) Heatmap showing module-MRI outcomes associations for BPF, WMF, GMF, and CTH. The x-axis indicates MS outcomes, and the y-axis indicates WGCNA-derived metabolite modules. Statistical significance is indicated as *p<0.05; **FDR<0.05. The adjacent stacked bar plot shows the metabolites class composition and metabolite count for each module; colors indicate metabolite classes. (B) Volcano plots showing associations between baseline individual metabolites and longitudinal MS outcomes, metabolites with FDR<0.05 are labeled.

**Table 2.**
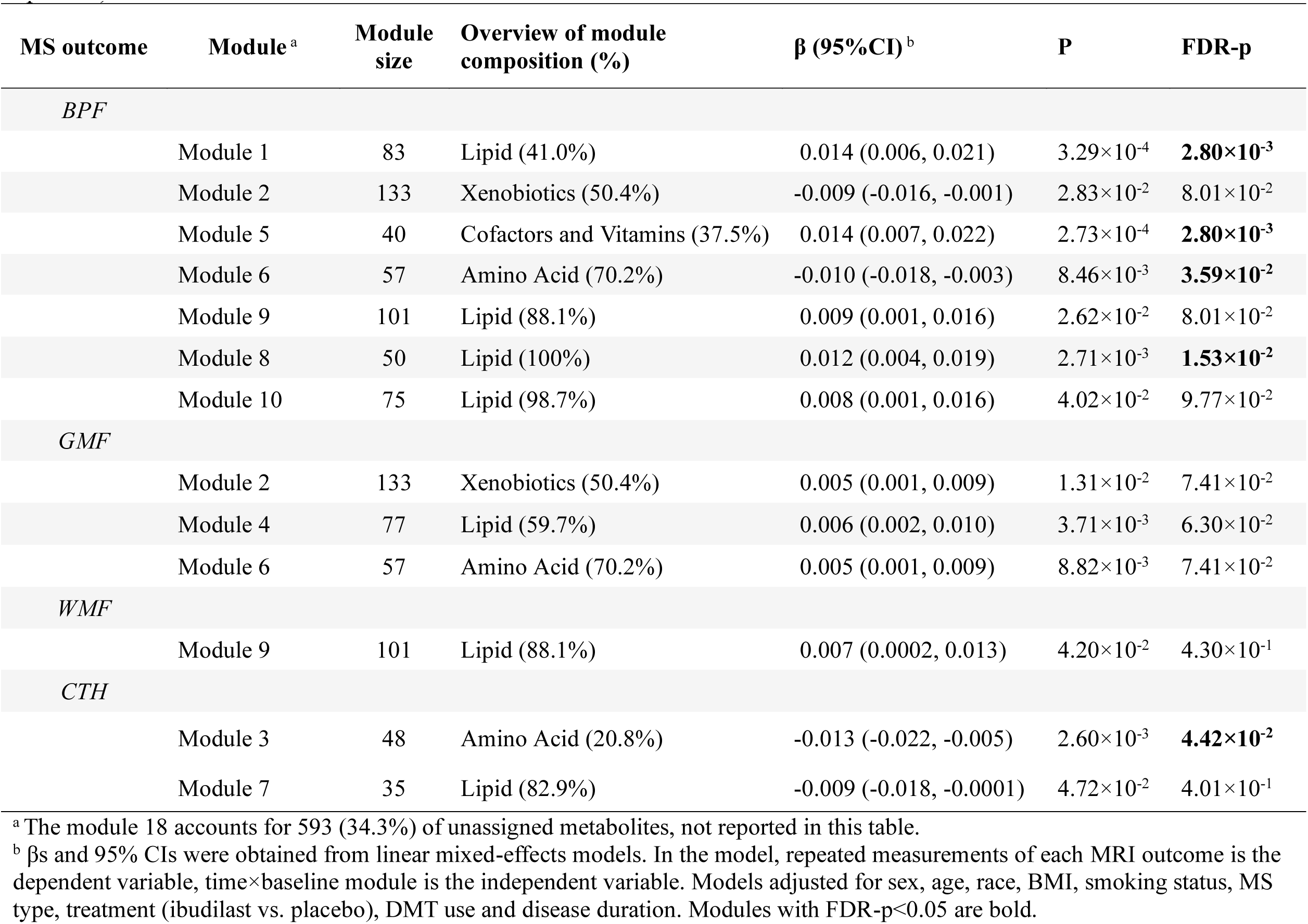
Baseline metabolite modules associated with longitudinal brain atrophy outcomes in MS in SPRINT-MS (list all 10 modules at p<0.05).

At the individual-metabolite level, the MRI-associated modules showed largely consistent metabolite-level patterns that corroborated the module-level findings. Specifically, for BPF, 204 metabolites (186 named) were associated with longitudinal change at p<0.05. Most associations were in the protective direction (144 out of 204 metabolites) and were predominantly glycerophospholipids and sphingomyelins (**Figure 2B**). Among the 180 BPF-related metabolites measured in HEAL-MS (**Supplemental Table S2**), 1-palmityl-2-stearoyl-GPC (O-16:0/18:0) was associated with slower BPF decline in SPRINT-MS (β=0.016 [0.008, 0.024]; p=4.35×10⁻^5^) and replicated in HEAL-MS (β=0.108 [0.006, 0.211], p=3.90×10⁻^2^). In contrast, homoarginine (β =−0.009 [−0.017 to −0.001]; p=2.08×10⁻^2^) and picolinoylglycine (β=−0.010 [−0.018 to −0.003]; p=7.37×10⁻^3^) were associated with greater BPF decline in SPRINT-MS and replicated in HEAL-MS (β =−0.111 [−0.202 to −0.020], p=1.79×10⁻^2^; and β =−0.143 [−0.271 to −0.015], p=2.92×10⁻^2^, separately) (**Figure 3**).

**Figure 3.**
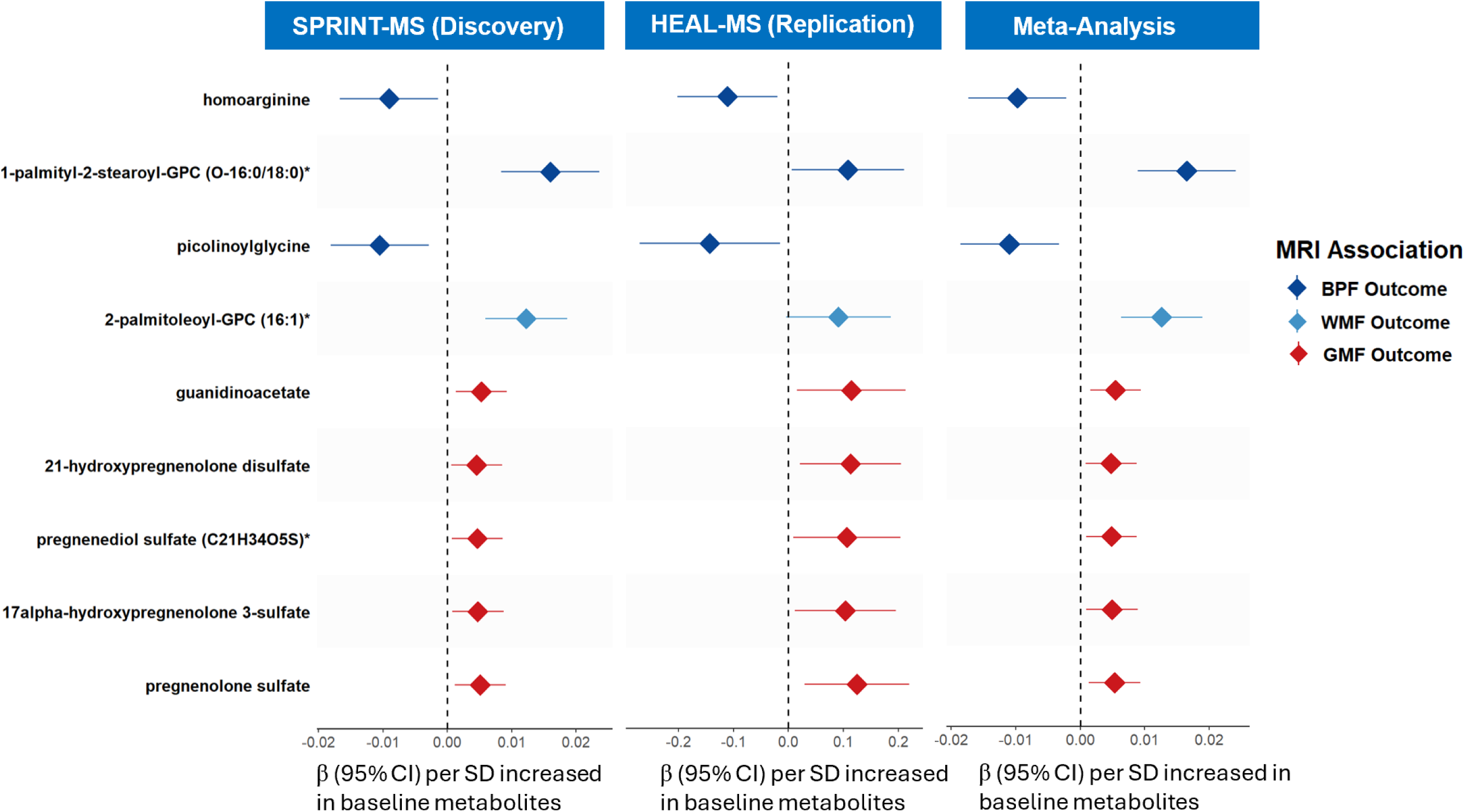
Baseline metabolites associated with longitudinal BPF, WMF or GMF in SPRINT-MS (discovery) and HEAL-MS (replication). Regression coefficients (βs) and 95% CIs in SPRINT-MS were obtained by linear mixed-effects models, adjusting for age, sex, race, BMI, smoking status, treatment group, MS subtype, disease duration, and baseline DMT use. βs in HEAL-MS (replication) were obtained by a linear mixed-effects model, adjusting for age, sex, race, BMI, smoking status, MS subtype, disease duration, and baseline DMT use. Diamonds are color-coded by MRI outcome: BPF (dark blue), WMF (light blue), and GMF (red).

For WMF, 27 metabolites (all named) were associated with longitudinal change at p<0.05, most of which were glycerophospholipids (**Figure 2B**). Among the 25 WMF-associated metabolites also measured in HEAL-MS, 2-palmitoleoyl-GPC (16:1) showed a consistent positive association that approached nominal significance in HEAL-MS (β=0.091[−0.004, 0.186]; p=6.24×10⁻^2^) (**Figure 3; Supplemental Table S3**).

For GMF, 100 metabolites (75 named) were associated with longitudinal change at p<0.05 (β range: 0.004 [0.001, 0.008] to 0.006 [0.003, 0.011]). Approximately half were androgenic steroids or steroid sulfates (**Figure 2B**). Of the 92 GMF-related metabolites also measured in HEAL-MS (**Supplemental Table S4**), five metabolites replicated with consistent positive associations, including guanidinoacetate and four steroid-related metabolites (i.e., 21-hydroxypregnenolone disulfate, 17-alpha-hydroxypregnenolone 3-sulfate, pregnenolone sulfate, and pregnanediol sulfate (C21H34O5S)) (β range: 0.104 [0.012, 0.196] to 0.125 [0.030, 0.220]; all p<0.05) (**Figure 3**).

For CTH, 24 metabolites (17 named) were associated with longitudinal change at p<0.05, of which 22 showed inverse associations (**Figure 2B**, **Supplemental Table S5**). After FDR correction, eight metabolites (4 named: p-cresol sulfate, 3-indoxyl sulfate, 11-ketoetiocholanolone glucuronide, 11 beta-hydroxyetiocholanolone glucuronide) remained significantly associated with greater CTH decline (β range: −0.018 [−0.026 to −0.009] to −0.012 [−0.020 to −0.004]). In contrast, two metabolites (dextromethorphan, 6-sulfatoxymelatonin) were associated with relative CTH preservation (β range: 0.012 [0.003, 0.020] to 0.018 [0.009, 0.026]; FDR<0.05).

### Ibudilast Association with Longitudinal Trajectories of Metabolites in SPRINT-MS

Across the 18 WGCNA-derived metabolite modules, three showed differential longitudinal trajectories between the ibudilast and placebo groups at p<0.05 (**Table 3**). Compared with placebo, ibudilast treatment was associated with an increase in the sphingolipid-enriched module over time (module 8; β=0.117 [0.018, 0.216]; p=2.12×10⁻^2^), and decreases in the xenobiotic-enriched module (module 3; β=−0.127 [−0.243 to −0.010]; p=3.36×10⁻^2^) and steroid-enriched module (module 4; β = −0.087 [−0.159 to −0.015]; p=1.84×10^−2^). Notably, these treatment-associated modules overlapped with modules implicated in MRI outcomes: higher baseline levels of sphingolipid-enriched module (module 8) were associated with slower BPF decline, whereas xenobiotic-enriched module (module 3) was associated with greater cortical thinning.

**Table 3.** Longitudinal changes in metabolite modules associated with ibudilast treatment identified by WGCNA in SPRINT-MS (list 3 modules at P<0.05).

| Module<sup>a</sup> | Module size | Overview of module composition (%) | $\beta$ (95%CI)<sup>b</sup> | P value | FDR-P |
| --- | --- | --- | --- | --- | --- |
| Module 3 | 48 | Amino Acid (20.8%) | -0.127 (-0.243, -0.010) | $3.36 \times 10^{-2}$ | $2.02 \times 10^{-1}$ |
| Module 4 | 46 | Amino Acid (69.6%) | -0.087 (-0.159, -0.015) | $1.84 \times 10^{-2}$ | $1.91 \times 10^{-1}$ |
| Module 8 | 50 | Lipid (100%) | 0.117 (0.018, 0.216) | $2.12 \times 10^{-2}$ | $1.91 \times 10^{-1}$ |

<sup>a</sup> The module 18 accounts for 593 (34.3%) of unassigned metabolites, not reported in this table.
<sup>b</sup> βs and 95% CIs were obtained from linear mixed-effects models. In the model, repeated measurements of each module is the dependent variable, time × treatment (ibudilast vs. placebo) is the independent variable. Models adjusted for sex, age, race, BMI, smoking status, MS subtype, DMT use and disease duration at baseline.

At the individual metabolite level, 13 metabolites within the sphingolipid-enriched module (module 8) increased over time with ibudilast treatment at p<0.05 (**Figure 4A**), most of which were sphingomyelins. Among these, palmitoyl sphingomyelin (d18:1/16:0) remained significant after multiple testing correction (β=0.185 [0.085, 0.286]; FDR=1.79×10^−2^) (**Figure 4B**). In contrast, 29 metabolites in the steroid-enriched module (module 4) and 10 metabolites in the xenobiotic-enriched module (module 3) decreased over time with ibudilast treatment at p<0.05. Most metabolites in module 4 were androgenic steroids, and 22 remained significant after FDR correction. In the xenobiotic-enriched module (module 3), anthranilate was the only metabolite that remained significantly decreased after FDR correction (β=−0.270 [−0.403, −0.137]; FDR=3.87×10^−2^). A complete list of significant ibudilast–metabolite associations is provided in **Supplementary Table S6**.

**Figure 4.**
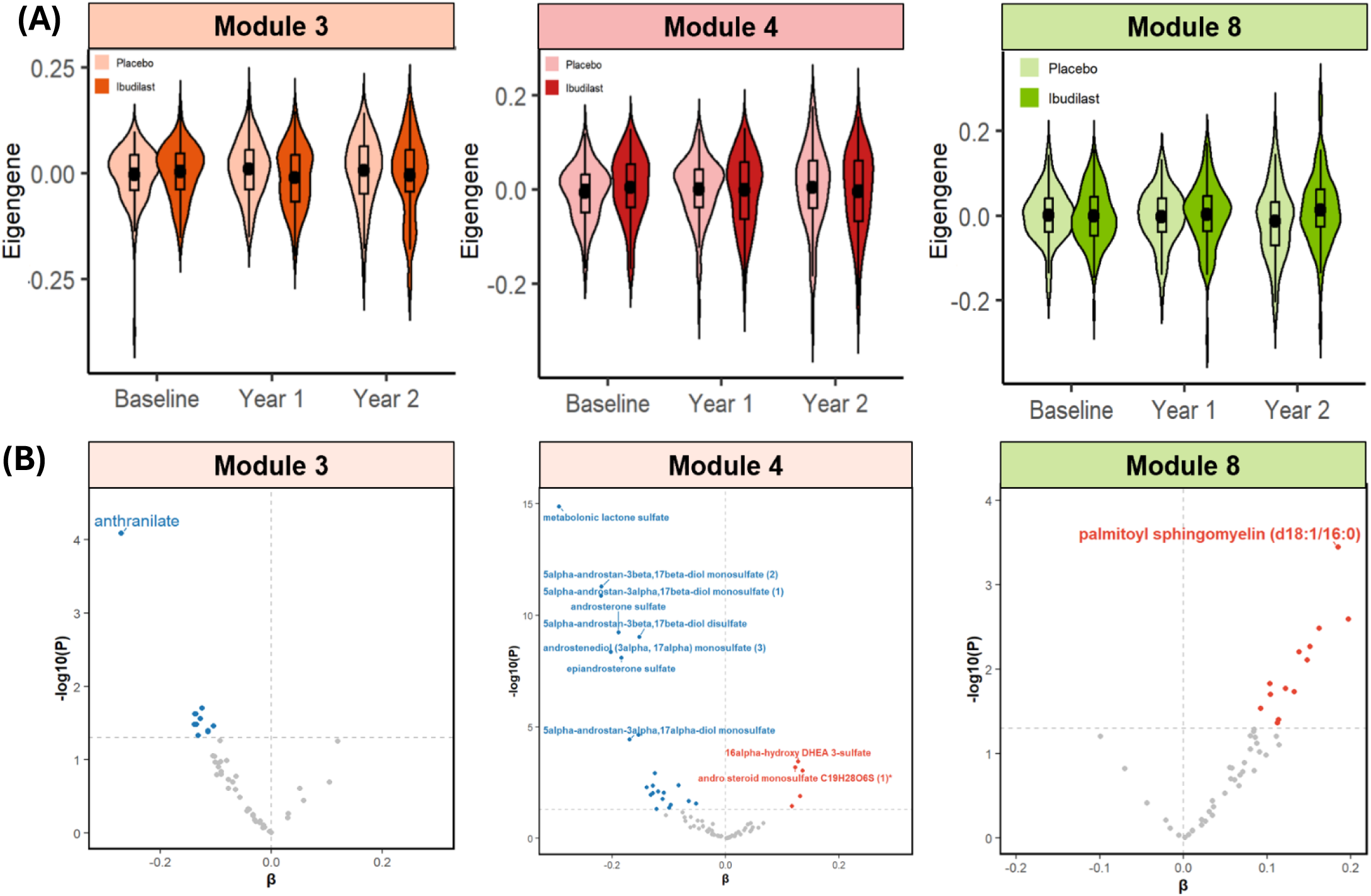
Longitudinal trajectories of metabolite modules and constituent metabolites associated with ibudilast treatment in SPRINT-MS. (A) Violin plots showing the distribution of module eigenmetabolites (z-scores) at baseline, year 1, and year 2 for participants randomized to ibudilast vs placebo. Panels display module 8 (sphingolipid-enriched module), module 3 (xenobiotic- and gut microbial metabolite-enriched module), and module 4 (androgenic steroid and steroid sulfate-enriched module), each showing a significant ibudilast-related change over time (p<0.05). (B) Volcano plots showing the ibudilast-associated changes in individual metabolites within each module, estimated using linear mixed-effects models adjusted for prespecified covariates; Each point represents a metabolite. Metabolites with FDR <0.05 are labeled.

### Metabolite Set Enrichment Analysis

MSEA identified outcome-specific metabolic pathways across MRI measures of brain atrophy (**Figure 5; Supplementary Table S7**). For BPF, lipid-related pathways, including phosphatidylcholine, lysophospholipid, long chain monounsaturated fatty acid, sphingomyelin, and dihydrosphingomyelin metabolism, were positively enriched. In contrast, amino acid-related pathways, including urea cycle/arginine and proline metabolism and gamma-glutamyl amino acids, as well as androgenic steroid metabolism, were inversely enriched. For GMF, steroid-related pathways, including androgenic and pregnenolone metabolism, branched-chain amino acid metabolism, and xanthine metabolism, were positively enriched. In contrast, sphingomyelin and monoacylglycerol metabolism were inversely enriched. For WMF, xanthine metabolism was positively enriched. For CTH, androgenic steroids and acetylated peptides pathways were inversely enriched, whereas selected lipid-related pathways, including diacylglycerol and monoacylglycerol, were positively enriched. Because MSEA included all measured metabolites, these analyses provided pathway-level summaries beyond the module-restricted WGCNA findings.

**Figure 5.**
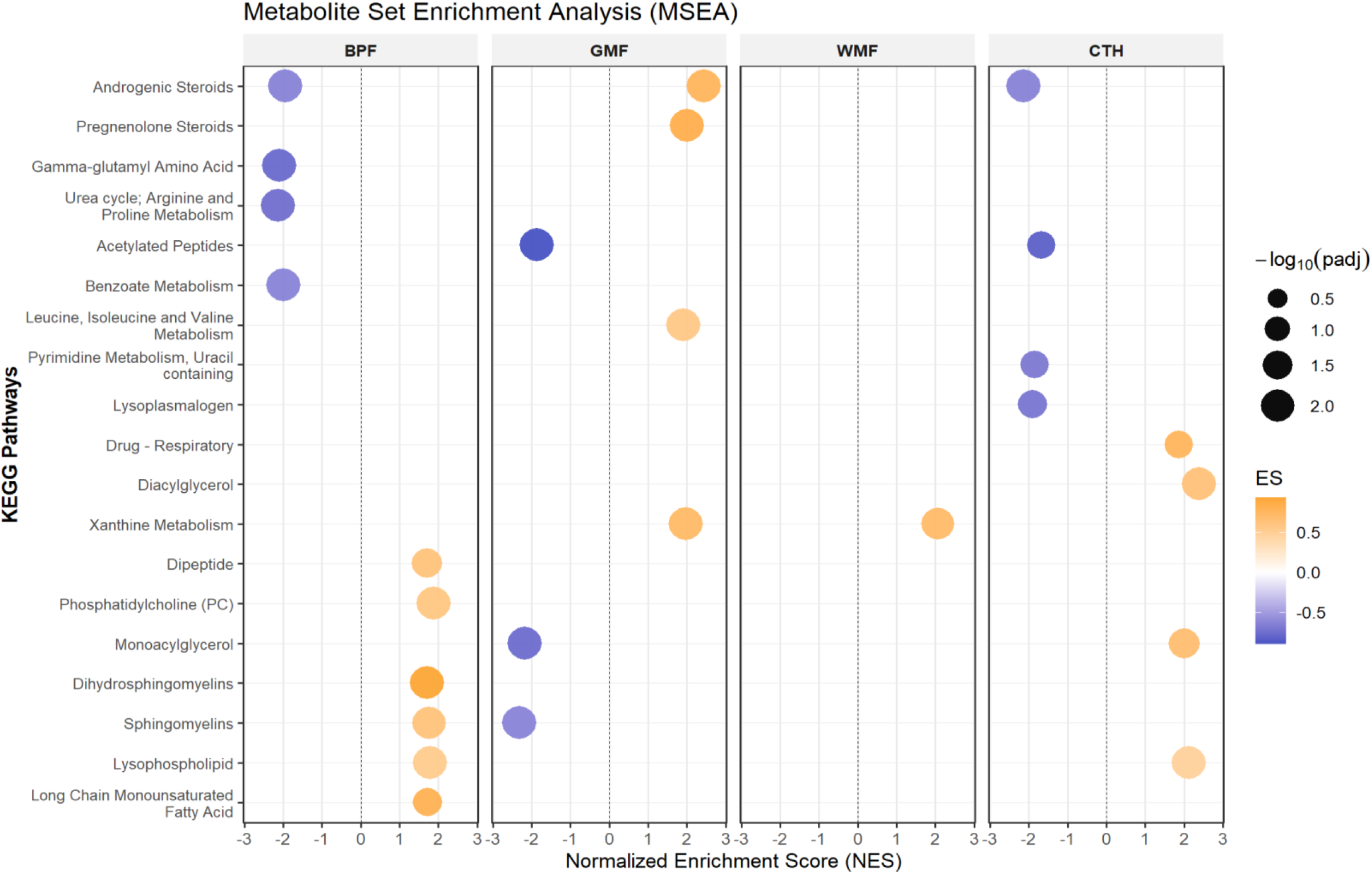
Biological pathways associated with MRI outcomes in SPRINT-MS. Dot plot showing MSEA results for BPF, GMF, WMF, and CTH. Pathways are displayed along the y-axis, and x-axis represents the normalized enrichment scores (NES), reflecting the degree of overrepresentation for each metabolite set within a ranked list of analyzed metabolites, adjusted for set size. Dot size corresponds to −log10 of the FDR-adjusted P value, indicating statistical significance. The color gradient represents the enrichment score (ES) of each pathway.

### Sensitivity Analysis

We evaluated sex-specific associations between baseline metabolites and longitudinal MRI outcomes in SPRINT-MS (**Supplementary Table S8**). Several hormone-related, xanthine-related, and xenobiotic metabolites showed nominal sex interactions for WMF, GMF, and CTH; however, most did not remain significant after correction for multiple testing. The strongest FDR-significant interaction was observed for 2-palmitoleoyl-GPC (16:1), for which higher baseline levels were associated with slower WMF decline in females compared with males (β=0.019 [0.006, 0.032]; FDR=1.94×10^−2^).

Given the enrichment of GMF-associated metabolites in steroidogenic or xanthine-related pathways, we also conducted sex-stratified analyses. In females, xanthine-related metabolites involved in caffeine metabolism, including caffeine, 1,3,7-trimethylurate, 1,7-dimethylurate, 1-methylxanthine, and 1-methylurate were positively associated with GMF and remained significant after multiple testing (all FDR < 0.05) (**Supplementary Table S8)**. No sex-specific effects were observed for associations between ibudilast treatment and longitudinal metabolite trajectories.

## Discussion

In this secondary analysis of a multicenter randomized clinical trial of progressive MS with external replication in an independent observational cohort, we identified several important findings. First, higher baseline levels of glycerophospholipids were associated with slower decline in both BPF and WMF, while sphingomyelins, another dominant lipid class, were similarly linked to slower BPF decline. In contrast, higher baseline levels of androgenic steroids were associated with slower GMF decline, whereas a subset of xenobiotic-related metabolites was associated with faster CTH decline. Several key metabolites were independently replicated in an external cohort of similar age distribution. Second, ibudilast treatment was associated with longitudinal metabolic changes that paralleled these observational patterns, including increases in sphingomyelin-enriched modules and higher levels of specific sphingomyelin species such as palmitoyl sphingomyelin (d18:1/16:0), alongside decreases in xenobiotic-enriched modules and lower levels of metabolites such as anthranilate. Finally, pathway enrichment analyses identified outcome-specific metabolic pathways across MRI measures of brain atrophy and highlighted dysregulation of lipid metabolism as a key pathway implicated in MS-related brain atrophy.

We identified higher baseline levels of glycerophospholipids, particularly phosphatidylcholines (PCs) and lysophosphatidylcholines (lysoPCs), as the dominant signatures associated with slower decline in both BPF and WMF. Replication of key species (e.g., 1-palmityl-2-stearoyl-GPC (O-16:0/18:0)) in an independent cohort supports the robustness of these associations. These findings align with prior studies reporting reduced circulating PC levels in people with MS^51^ and associations between lower PC concentrations and greater disease severity.^52^ The positive association of 2-palmitoleoyl-GPC (16:1) with WMF preservation is also supported by Mendelian randomization studies supporting protective roles of related monounsaturated PC species in neurological disease,^53^ as well as epidemiologic evidence linking this metabolite to a lower risk of major depressive disorder.^54^ Interestingly, prior studies have linked 1-palmityl-2-stearoyl-GPC (O-16:0/18:0) to increased glioblastoma risk^55^ and Alzheimer’s disease-related pathology, ^56^ whereas in our study it was associated with greater BPF preservation. This discrepancy may reflect variations in biological compartment or disease-specific lipid metabolism. Further studies are needed to clarify its role in MS progression. Given the established role of phospholipids in membrane structure and myelin biology,^57–59^ these findings support a role for glycerophospholipid pathways in MS-related neurodegeneration, although the underlying mechanisms remain to be clarified.

We observed that higher baseline levels of androgenic steroids (e.g., androsterone glucuronide, epiandrosterone sulfate, and dehydroepiandrosterone sulfate [DHEA-S]) were associated with slower GMF decline. This is consistent with prior evidence demonstrating reduced circulating testosterone and DHEA levels in people with MS.^60^ Small interventional studies have further shown that testosterone supplementation in men with relapsing-remitting MS improved cognitive performance, slowed whole-brain atrophy,^61^ and increased gray matter volume.^62^ Our results extend this evidence by identifying specific sulfated and glucuronidated androgen metabolites associated with longitudinal GMF preservation, suggesting that conjugated steroid metabolism may serve as integrated markers of endocrine influences on brain atrophy. Sex hormones, including estrogens, progesterone, and androgens, modulate immune activation, remyelination, and neuroinflammatory signaling, processes central to MS pathogenesis.^63^ Experimental studies demonstrate that androgens promote oligodendrocyte differentiation, enhance myelin repair, and suppress microglial activation.^64^

Additionally, we found several metabolites (e.g., 3-indoxyl sulfate, 6-sulfatoxymelatonin, dextromethorphan) were associated with longitudinal changes in CTH. Higher baseline levels of 3-indoxyl sulfate were associated with greater cortical thinning, consistent with prior evidence linking this gut-derived uremic toxin to oxidative stress, endothelial dysfunction, and neuroinflammation, processes that can exacerbate brain atrophy.^65,66^ In contrast, 6-sulfatoxymelatonin, the principal metabolite of melatonin, was positively associated with preserved cortical structure. Altered 6-sulfatoxymelatonin levels have been reported in relapsing-remitting MS and are associated with greater disability and fatigue.^67^ This metabolite has been explored as a biomarker of treatment response to fluoxetine in major depressive disorder.^68^ Mechanistically, melatonin has been shown to modulate immune responses relevant to MS pathogenesis, including suppression of pathogenic Th17 cell differentiation and enhancement of regulatory Tr1 cells, potentially linking circadian and environmental factors to neuroinflammatory activity.^69^ These findings are consistent with the antioxidant, anti-inflammatory, and circadian-regulating functions of the melatonergic pathway in the CNS.^70^ Dextromethorphan, an NMDA- receptor antagonist with anti-glutamatergic and neuroprotective properties,^71^ was also associated with slower CTH decline; although exogenous, its presence may reflect medication exposure and should be interpreted cautiously. Future studies are warranted to replicate these metabolites in independent cohorts.

Importantly, ibudilast treatment was associated with metabolic changes that paralleled baseline signatures of brain atrophy, including increases in sphingomyelin-related pathways and reductions in tryptophan-kynurenine metabolites. This pattern is notable because higher baseline levels of sphingomyelin were associated with slower BPF decline, suggesting that treatment-related increases may reflect modulation of lipid pathways linked to membrane integrity and myelin stability. Sphingomyelins are key components of myelin and play important roles in neuroinflammatory signaling, and their dysregulation has been implicated in MS pathophysiology.^72,73^ Prior studies have reported reduced circulating and tissue sphingomyelin species in MS,^74^ as well as associations between higher long-chain sphingomyelin levels and preserved white matter integrity in longitudinal studies.^75^ Experimental evidence further supports the relevance of sphingolipid signaling in demyelination and disease severity, including pathways involving sphingosine-1-phosphate receptors.^76^ In contrast, ibudilast was associated with reductions in tryptophan-kynurenine pathway metabolites, including anthranilate. This pathway is central to immune regulation and neuroinflammation, and its dysregulation has been reported in MS^77^ and linked to inflammatory shifts toward neurotoxic metabolites, disease activity and neurodegeneration.^78^

Another notable finding was that pathway enrichment analysis revealed divergent associations across MRI measures of brain atrophy. Androgenic steroid pathways showed positive enrichment for GMF but inverse enrichment for BPF, whereas lipid-related pathways, particularly sphingomyelins, showed positive enrichment for BPF but inverse enrichment for GMF. These patterns may reflect outcome-specific biological information captured by different neuroimaging metrics rather than uniform pathway effects across all measures of atrophy. BPF reflects global parenchymal tissue loss across gray and white matter compartments, whereas GMF more specifically reflects gray matter volume, including cortical and deep gray matter structures. The positive enrichment of androgenic steroid pathways for GMF is consistent with prior evidence linking androgen signaling to gray matter structure^79^ and with potential neuroprotective effects in MS.^64^ By comparison, the divergent associations observed for lipid-related pathways may reflect compartment-specific differences in lipid biology. Myelin and white matter are substantially more lipid-rich than gray matter, and classic lipidomic studies have demonstrated marked differences in lipid composition among myelin, white matter, and gray matter.^80^ Given the indirect nature of MRI-derived measures and the use of circulating metabolites, our findings should be considered hypothesis-generating and warrant validation using CSF lipidomics, tissue-based studies, and multimodal imaging approaches to clarify compartment-specific metabolic mechanisms in MS.

Several limitations of this study should be noted. Although the sample size of SPRINT-MS was relatively large for a progressive MS clinical trial, statistical power remained limited to detect small metabolite effects after multiple-testing correction. The generalizability of our findings may also be constrained by the study populations. SPRINT-MS enrolled only participants with progressive forms of MS, more than half of whom had primary progressive MS. While key findings were replicated in the independent HEAL-MS cohort, additional validation is needed in broader, younger MS populations, including earlier relapsing-onset disease and more racially and clinically diverse settings. Moreover, blood-based metabolomics captures systemic metabolic signatures that may not fully reflect CNS-specific processes underlying brain atrophy. Fasting status at the time of blood collection was not recorded in SPRINT-MS; therefore, residual effects of recent dietary intake on circulating metabolites cannot be excluded. Nevertheless, major metabolite classes identified in this study, particularly sphingomyelins and glycerophospholipids, have demonstrated relatively high intra-individual stability^81^ and minimal short-term variability under routine fasting and postprandial conditions.^82^ Although statistical models adjusted for a comprehensive set of demographic and clinical covariates, residual confounding from unmeasured factors cannot be fully ruled out. Finally, despite standardized MRI acquisition protocols and centralized imaging processing, minor scanner-related variability across sites cannot be entirely ruled out.

The major strength of this study is its randomized, placebo-controlled design of the discovery cohort, one of the first studies to integrate metabolomics within a rigorously conducted clinical trial of progressive MS. This design minimizes confounding and enables standardized, high-quality collection of clinical, imaging, and biospecimen data. Another key strength is the longitudinal design, which included repeated MRI and plasma metabolomic assessments over 96 weeks, enabling evaluation of both metabolite trajectories and MRI-based measures of brain atrophy, as well as the effects of ibudilast treatment on metabolic pathways. Importantly, our findings were supported by replication in an independent cohort, comprising a total sample of over 490 participants across two cohorts. Several metabolite-MRI associations showed consistent directionality, reinforcing the robustness and generalizability of our results. Additionally, our statistical models were comprehensively adjusted for demographic and clinical factors, with sensitivity analyses further examining sex-specific associations. We also combined network analysis and pathway enrichment analyses to provide biologically coherent interpretations of the observed metabolic signatures. Finally, high-resolution untargeted metabolomic profiling captured a broad range of lipid and amino acid species, facilitating the identification of novel metabolites potentially involved in MS progression.

In summary, we identified distinct metabolic signatures associated with longitudinal MRI outcomes in progressive MS. Higher baseline levels of glycerophospholipids and sphingomyelins were consistently linked to a slower rate of brain atrophy, independent of clinical risk factors, and ibudilast treatment was associated with related metabolites in directions consistent with neuroprotection. These findings advance understanding of lipid dysregulation in progressive MS and highlight the potential of metabolomics to identify prognostic and pharmacodynamic biomarkers that could inform future therapeutic development.

## Data Availability

The SPRINT-MS and HEAL-MS clinical datasets are not publicly available; however, access requests can be submitted to the corresponding author.

## Contributors

KF conceptualized and designed the study, obtained the funding, and generated the data. MC conducted the statistical analysis. MC drafted the manuscript. KF, RN, CT, and PB helped with data interpretation and provided critical review and contributed to the revision of the manuscript. All authors read and approved the final version of the manuscript.

## Acknowledgements

The authors thank participants in SPRINT-MS and HEAL-MS.

