## Supplemental methods for "Metabolomic Signatures of Brain Atrophy and Ibudilast Response in Progressive Multiple Sclerosis"

Mingjing Chen,^1^ Rose Noroozi,^1^ Matthew D. Smith,^1^ Muraleetharan Sanjayan,^1^ Cesar Higgins Tejera,^1^ Pavan Bhargava,^1^ Blake E. Dewey,^1^ Ellen M. Mowry,^1^ Kathryn C. Fitzgerald^1,2*^

^1^Department of Neurology, Johns Hopkins University School of Medicine, Baltimore, MD, USA.

^2^Department of Epidemiology, Johns Hopkins University, Baltimore, MD, USA.

**Correspondence to**: Kathryn C. Fitzgerald, ScD.

Department of Neurology, Johns Hopkins University School of Medicine

Baltimore, MD, USA, 33612

**Online Supplementary Materials**

**Supplemental Methods.** Weighted Gene Co-expression Network Analysis (WGCNA) adopted for metabolomics: Soft-Threshold Power and Module Dendrogram.

**Supplemental Methods.** Pathways Enrichment Analysis for Individual Metabolites

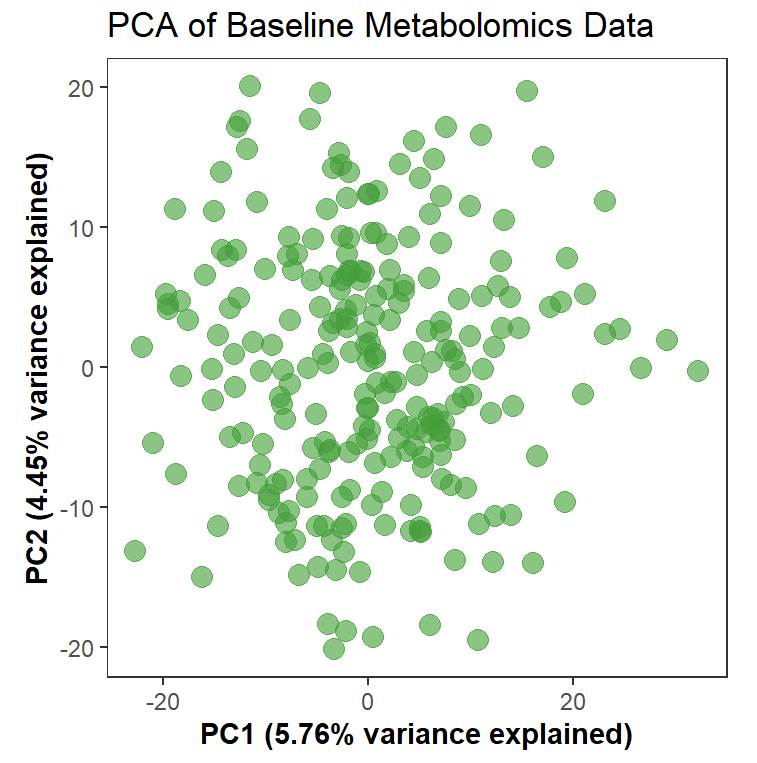
**Supplemental Methods.** Weighted Gene Co-expression Network Analysis (WGCNA) adopted for metabolomics in SPRINT-MS: Soft-Threshold Power and Module Dendrogram. An expression matrix of baseline samples from 244 participants was used to construct weighted co-expression networks for 1,726 metabolites using the WGCNA R package.^1^ Pairwise Pearson correlations were first calculated to generate a co-expression matrix. The “pickSoftThreshold” function was then used to identify an appropriate soft-thresholding power, chosen based on achieving a scale-free topology fit index above 0.85 while maintaining adequate mean connectivity. The resulting adjacency matrix was transformed into the topological overlap matrix (TOM), and modules were identified using hierarchical clustering with the dynamic tree cut method. The minimal module size was set to 30 for metabolites. Modules with similar expression profiles were merged using a height cutoff of 0.25. Module eigenmetabolites (MEs), defined by the first principal component of each module, were computed to summarize expression patterns. (A): PCA of baseline metabolomics data after quality control (B): Soft-thresholding power selection for baseline metabolomics data (C): Metabolites dendrogram and module colors

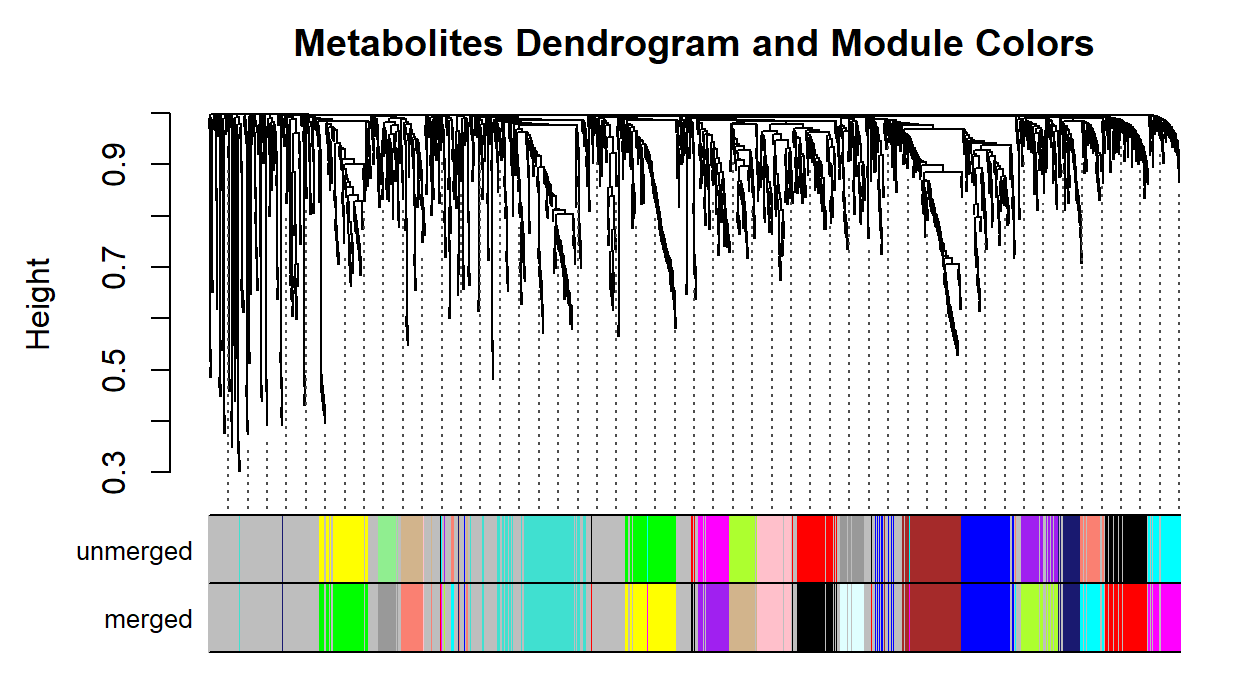

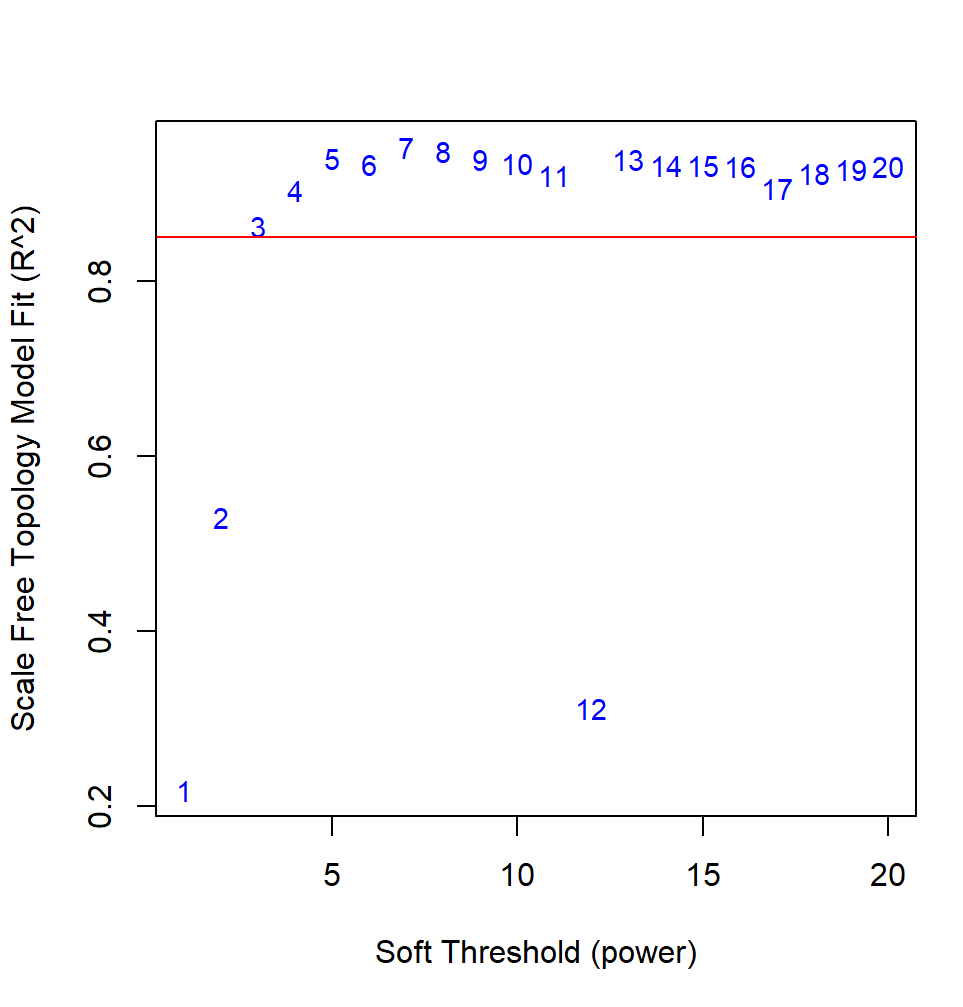

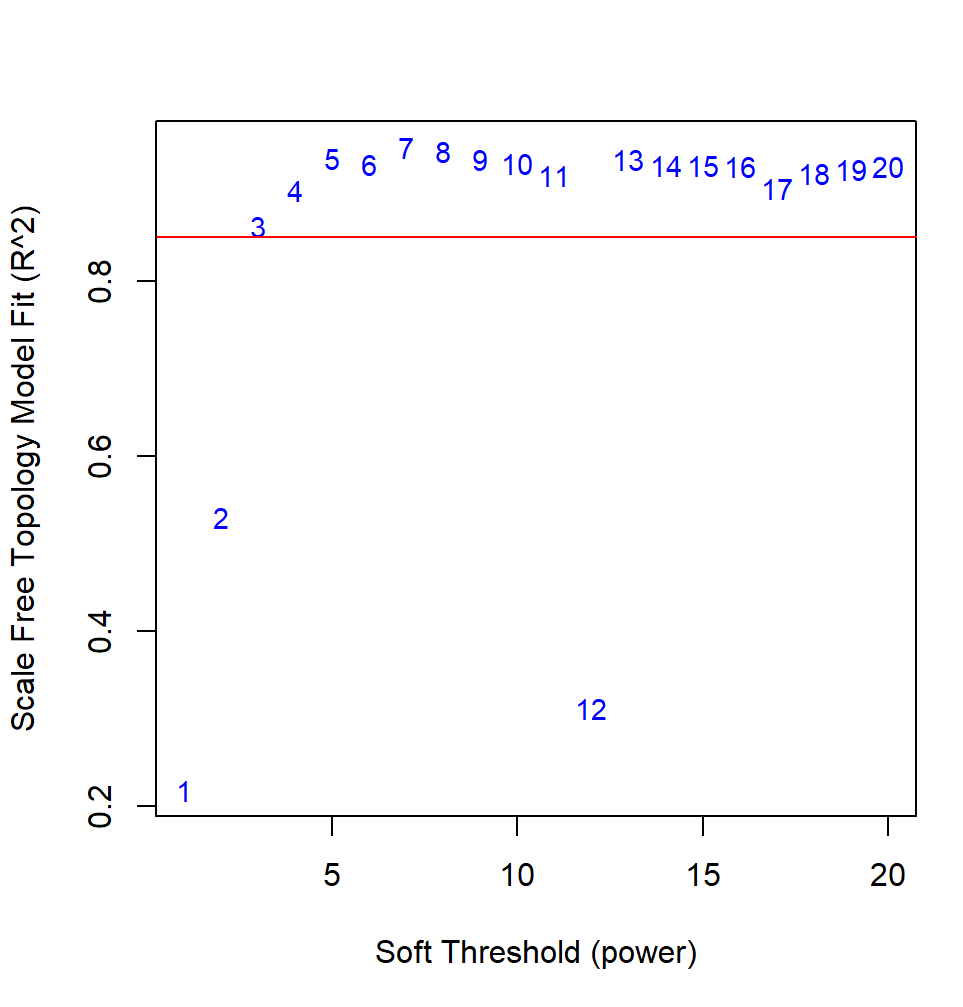

**A**

**B**

**C**

**Supplemental Methods.** Pathways Enrichment Analysis for Individual Metabolites

Pathway enrichment analysis was performed using a metabolite set enrichment analysis (MSEA) framework based on predefined metabolic sub-pathways provided by the Metabolon platform. Metabolites were annotated according to Metabolon-defined super-pathways and sub-pathways, which are curated based on biochemical classification and known metabolic relationships.

For each MRI outcome, metabolites were ranked according to their test statistics derived from the primary association analyses. These ranked lists were used as input for enrichment analysis to evaluate whether metabolites belonging to a given sub-pathway were disproportionately represented at the extremes of the ranked distribution.

Enrichment scores (ES) were calculated to quantify the degree to which each metabolite set was overrepresented among the most positively or negatively associated metabolites. To account for differences in pathway size, normalized enrichment scores (NES) were computed, enabling comparisons across metabolite sets of varying sizes.

Statistical significance was assessed using permutation-based testing, in which metabolite labels were permuted to generate an empirical null distribution of NES values.^3,4^ Resulting p-values were adjusted for multiple comparisons using the false discovery rate (FDR) method. Pathways with FDR-adjusted p-values below the predefined significance threshold were considered significantly enriched.
